# A Blood test to measure proteins for a multi-cancer diagnostic

**DOI:** 10.1101/2025.05.07.25326455

**Authors:** Douglas D. Held, Steven Bolland, Robert Freese, Robert S. Puskas

## Abstract

Current cancer screening is primarily restricted to various forms of imaging. For breast and lung cancer screening there are a significant percentage of false positive tests. Testing for colorectal involves invasive colonoscopies. For some of these tests, recent recommendations to extend the screening age from 45 to 40 put more pressure on a screening system that already has difficulty meeting the suggested recommendations for screening – especially in underserved rural areas. There is a need for a simple effective screening method that can test the recommended population. Samples (154 cancer and 119 normals) were tested for five different cancers (breast, lung, colorectal, ovarian, and pancreatic). The test included measurement of extracellular protein kinase activity, other kinase activities, and antibodies to altered cancer proteins (both IgG and IgM forms). The resulting test had a sensitivity of 100% with an overall specificity of 97%. Importantly, the test also identified the cancer tissue of origin (TOO) 98% of the time minimizing the need for downstream confirmatory testing and biopsies. The test had the added benefit of differentiating cancer subtypes for lung and ovarian cancers. The groundwork was established for an effective multi-cancer early detection (MCED) test for five different cancers

## Introduction

Screening for cancer has multiple shortcomings. Imaging for screening (mammograms for breast cancer and LD-CAT scans for lung cancer) has high false positives. Ten percent of mammograms are apparent positives with 95% of these being false positives while 33% of LD-CAT scans are false positives (1, 2). Mid-sized nodules detected by LD-CAT scans are confirmed as cancer by measuring doubling times measured by repeated LD-CAT scans at 3 to 6-month intervals for up to a year (3). However, some subtypes of lung cancer can double in size in 3 months (4). Furthermore, compliance with testing intervals is far from perfect with 78% of women receiving mammograms, 60% of eligible individuals receiving colonoscopies, and less than 10% of eligible individuals receiving LD-CAT scans at recommended intervals (5-7). In addition, some cancers such as ovarian and pancreatic cancers have no recommended screening tests.

Nonetheless, female breast cancer mortality has decreased by 11% because of earlier diagnosis via screening. In contrast, female breast cancer incidence rates have been increasing with the steepest rate increase in women under 50. Additionally, those younger than 50 years were the only age group where cancer incidence increased from 1995 through 2021. Low-dose CT screening reduces lung cancer mortality between 16-24% in high-risk individuals (8). More reasons to provide simple screening procedures that encourage testing especially at younger ages.

One approach to resolving cancer screening shortcomings is liquid biopsies – blood tests that detect multiple cancers (MCDs). Blood tests are easy to administer and would lead to better testing compliance. However, these tests also have issues. Early detection of cancer is desirable as it generally leads to better patient outcomes. But current MCDs are unable to satisfy this requirement (9). An optimal MCD test also would have low false positives, low false negatives, and be able to efficiently identify the cancer tissue of origin (TOO). Current MCD tests often use circulating tumor DNA (ctDNA) or targets in cell-free DNA (cfDNA) to detect cancer. However, the concentrations of these target entities are low and difficult to detect at early cancer stages. Consequently, although tests like those from Grail (Galleri) are touted to detect 50 different cancers, many of them are detected at low sensitivities (9, 10). The MCD test from Thrive (Exact Sciences) (CancerSeek) suffers from the same issue (11). Both of these sources also have challenges determining the tissue of origin, with Grail leaving the physician to test for two or more types of cancer 1 in 15 times – this ratio is significantly higher for Thrive (9, 11).

The current study is focused on measuring abundant serum proteins that are found at the earliest stages of five different cancers, and which result in both high sensitivity, specificity and tissue of origin designation.

## Methods

Deidentified serum samples were purchased from commercial sources (ProMedDx and ProteoGenex). All patients consented to use of their samples for research.

Blood proteins measured in serum included extracellular protein kinase A (xPKA) activity, other kinases, and antibodies to altered cancer proteins. The MESACUP Protein Kinase Assay Kit (MBL, Product code 5230) was used to measure extracellular PKA in serum samples. A 108 µL serum sample was first mixed with 12 µL activating buffer (25 mM KH_2_PO_4_, 5mM EDTA, 150 mM NaCL, 50% glycerol (w/v), 1 mg/ml BSA, and 100 mM DTT, pH 6.5) and incubated at room temperature for 30 min to maximally activate the PKA. The activated xPKA (108 µL) was mixed with 108 µL of kit reaction buffer with or without 0.5 µM PKI, a protein kinase A inhibitor (Santa Cruz product sc-201160). A defined peptide bound to the wells of a microtiter plate was used as a substrate. The assay mixture was incubated in the presence of peptide for 30 min at 25°C with shaking at 750 rpm. Phosphorylation of the peptide was detected using biotinylated phosphoserine antibody per kit instructions, which was in turn was detected in an ELISA format using peroxidase conjugated to streptavidin. Detection of the bound peroxidase was established using a color-producing peroxidase substrate, TMB (Millipore Sigma T0440). TMB (100 µL) was directly added to all wells with a 60-min incubation/shake. Stop Solution (100 µL, 0.2M H_2_SO_4_) was added to all wells with a 30 second shake, followed by OD readings at 450nm. Bovine PKA catalytic subunit was used at varying concentrations to develop a standard activity curve. Net xPKA in the samples was calculated as Net xPKA = Kinase (0 µM PKI) – Kinase (0.5 µM PKI).

All antibody tests (IgG and IgM) used the same protocol. The only difference was the substrate coating of the microtiter plates (Maxisorp F96-well polystyrene) which was carried out manually.

Plate Coating: Group one antigens: 100 µL of the antigen (0.5ug/mL) was added to each well and incubated for 60 min at room temperature. Contents were aspirated. Pierce Protein-free Blocker Buffer (Thermo, Product #37572) was added to all wells, followed by 60 min incubation. Plates were washed with Coating Wash Buffer (20 mM HEPES, 150 mM NaCl, 30 mM Sucrose), allowed to dry and sealed. Group two antigens: 350uL of the antigen (200 µg/mL) was added to all wells with a 60 min incubation. Plates were then washed with Coating Wash Buffer, allowed to dry and sealed.

Antibody Test Protocol: This was a four-step protocol with incubation/shake (60 min at RT; 750rpm), plate aspiration and wash with Assay Wash Buffer (10 mM Citrate, 150 mM NaCl, 0.1% Tween 20) in the first two steps. Step one: each serum sample was diluted x200 in Assay Buffer (10 mM Na_2_PO_4_, 150 mM NaCl, 1% Tween 20). Diluted samples (100 µL) were transferred to appropriate wells in a coated plate. The plate was incubated/shaken, aspirated and washed. Step two: detection antibody (100 µL, anti-IgG or anti-IgM, Jackson ImmunoResearch) was added to all wells. The plate was incubated/shaken, aspirated and washed. TMB (100 µL, Millipore Sigma T0440) was directly added to all wells with a 60-minute incubation/shake. Stop Solution (100 uL, 0.2 M H_2_SO_4_) was added to each well with a 30 second shake, followed by OD readings at 450nm.

A total of 16 parameters were measured. Patterns for each cancer based on the protein measurements were determined for each cancer independently. The sample distributions and distributions by age are shown in Table 1.

**Table 1:** Demographics and Disease Characteristics.

| Category | Description | Ovarian | Lung | Colorectal | Pancreatic | Breast | Normal |
| --- | --- | --- | --- | --- | --- | --- | --- |
| AGE | *n* | 13 | 61 | 13 | 11 | 61 | 90 |
|  | mean | 56.31 | 63.28 | 62.85 | 63.18 | 56.33 | 58.52 |
|  | std | 13.193 | 7.572 | 8.849 | 11.241 | 12.504 | 7.897 |
|  | median | 54.00 | 63.00 | 62.00 | 63.00 | 54.00 | 57.50 |
|  | min | 31.00 | 45.00 | 51.00 | 37.00 | 32.00 | 46.00 |
|  | max | 74.00 | 76.00 | 78.00 | 76.00 | 79.00 | 75.00 |
|  | 95% CI | [48-64] | [61-65] | [57-68] | [55-70] | [53-59] | [56-60] |
| Ethnicity | Caucasian | (13, 100) | (61, 100) | (13, 100) | (11, 100) | (61, 100) |  |
| Sex | Female | (13, 100) | (17, 27.8) | (8, 61.5) | (8, 72.7) | (59, 96.7) | (70, 77.7) |
|  | Male |  | (44, 72.1) | (5, 38.4) | (3, 23.2) | (2, 0.3) | (20, 22.2) |
| Stage | 0 |  |  |  |  | (4, 6.6) |  |
|  | I |  |  | (1, 7.7) |  | (10, 16.4) |  |
|  | IA |  | (17, 27.9) |  |  | (6, 9.8) |  |
|  | IB | (1, 7.7) | (22, 36.1) |  | (1, 9.1) |  |  |
|  | II |  |  | (4, 30.8) |  |  |  |
|  | IIA | (2, 15.4) | (1, 1.6) |  | (2, 18.2) | (19, 31.1) |  |
|  | IIB |  | (3, .9) |  | (1, 9.1) | (10, 16.4) |  |
|  | III |  |  | (4, 30.8) | (4, 36.3) |  |  |
|  | IIIA | (4, 30.8) | (9, 14.7) |  |  | (4, 0.6) |  |
|  | IIIB |  | (4, 6.56) |  |  |  |  |
|  | IIIC | (5, 38.5) |  |  |  | (1, 1.6) |  |
|  | IV | (1, 7.7) | (3, 4.9) | (4, 30.8) | (3, 27.2) |  |  |
|  | N/A |  | (2, 0.3) |  |  | (7, 11.5) |  |

## Analysis

Statistical analysis of the data was performed using SAS® Version 9.4 (SAS Institute, Cary, NC, USA). All the data were standardized in terms of variables, labels, and overall presentation to facilitate its conversion into SAS datasets. Descriptive statistics are displayed to provide an overview of the results. For continuous variables, descriptive statistics include the number of samples with available measurements (n), mean, standard deviation (SD), the median, minimum, and maximum.

For categorical variables (Table 1), the number and percentage of samples in each in each age group, disease category, and Stage is presented. Unless otherwise noted, the denominator for percentages is based on the number of samples included.

## Results

Patterns of assay values for each cancer type were developed de novo. No assumptions were made that patterns from one cancer would appear in another cancer. At this point it was determined that 15% of the cancers had a pattern that appeared in another cancer type. The patterns were reassessed to minimize these “crossovers”. The effect was a reduction in crossovers to 2% resulting in a tissue of origin (TOO) designation of 98% (Table 2.). Consequently, follow up testing would be needed in very few cases to determine which cancer was present when a cancer was found. 100% of Stage I cancers were detected.

**Table 2:**
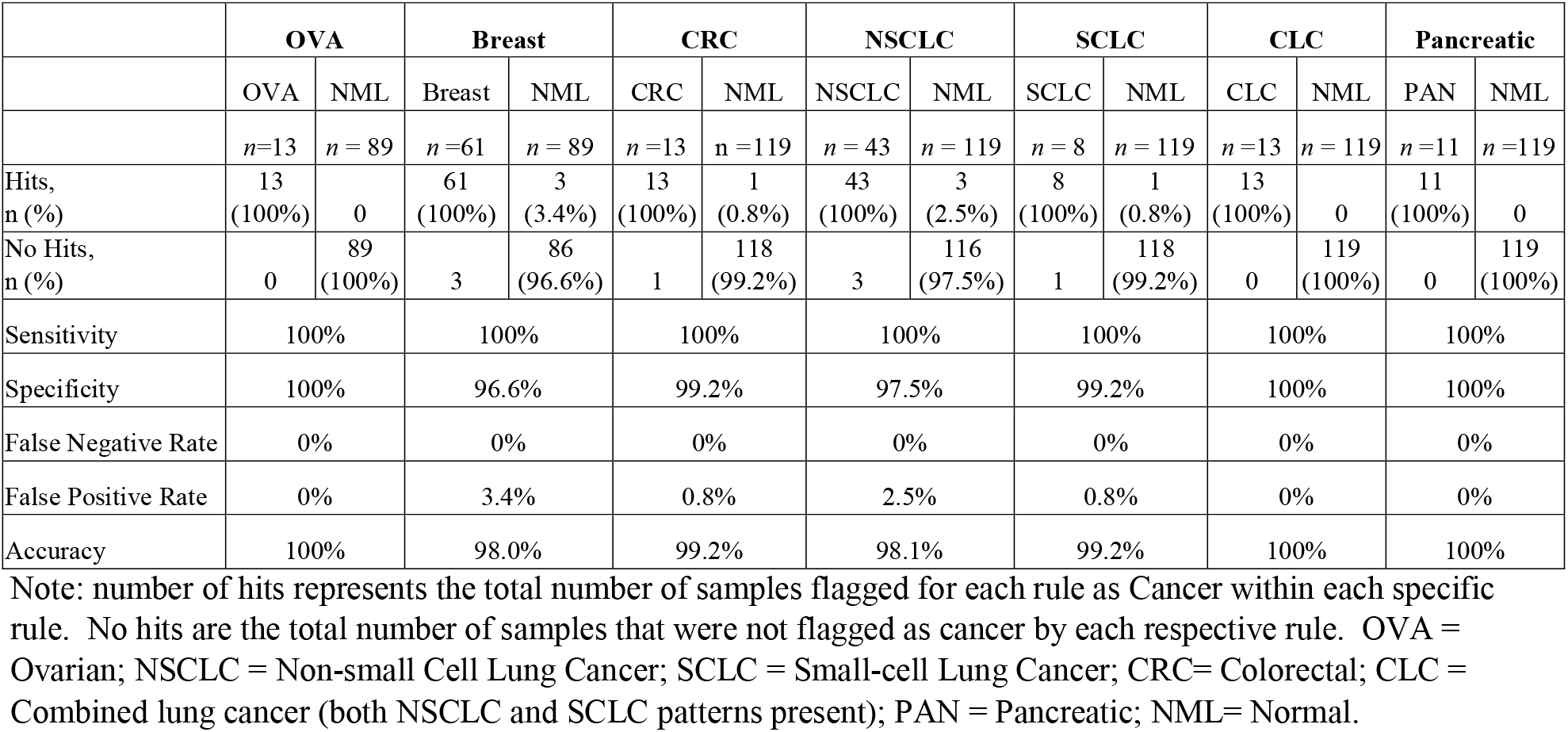
Overall Summary of Sensitivity, Specificity, False Negative Rate, False Positive Rate and Accuracy by Rule/Disease

Figure 1 shows the distribution of values for the key biomarker xPKA of the cancer samples relative to that of normal individuals. For most cancer types there is differentiation of the cancer values from those of the normals for this biomarker alone. The sensitivities and specificities of the MCD test are shown in Table 2 where the values are shown for each type of cancer. Sensitivities are 100% for each of the cancers and specificities range from 100% to 97%.

**Figure 1.**
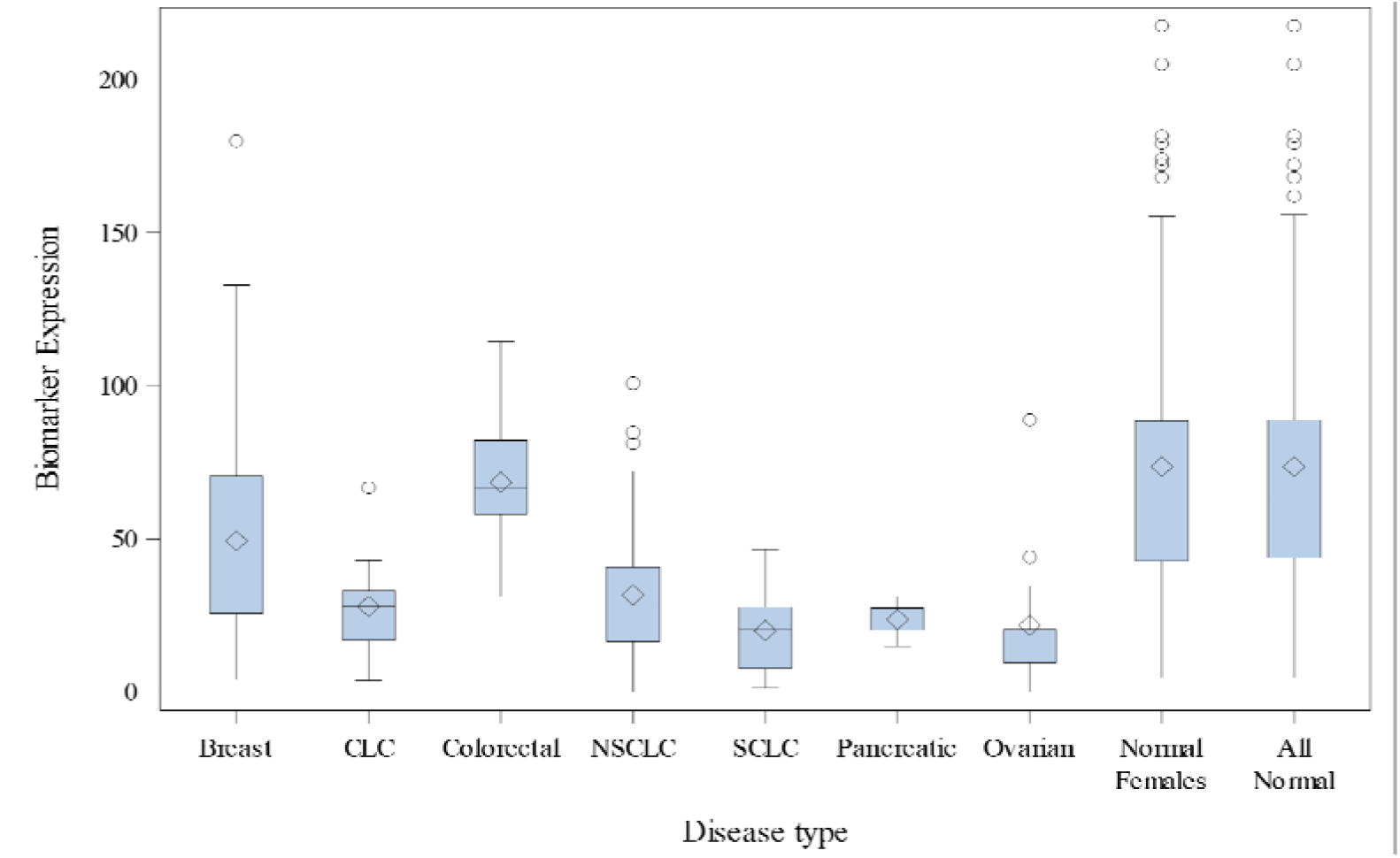
Box Plot for xPKA Activity by Disease Type Notes: The length of the box represents the interquartile range (the distance between the 25th and 75th percentiles) The diamond symbol in the box interior represents the group mean The horizontal line in the box interior represents the group median The vertical lines (called whiskers) issuing from the box extend to the group minimum and maximum values The empty circles represent outliers Program: boxplot.sas Alt Text. Distribution of values of extracellular protein kinase A activity for cancer samples and normals.

For lung cancer it was found that the aggressive SCLC subtype, found in about 15% of cases, could be distinguished from the more common NSCLC subtype. This is significant because SCLC is aggressive and benefits from different treatment than NSCLC. Furthermore, some NSCLC samples also had SCLC patterns indicating that these samples may have a mixed histology – both SCLC and NSCLC present.

In further differentiation, it was determined for ovarian cancer that the serous subtype could be distinguished from the mucinous subtype. More aggressive cancer is found among the serous subtype. Table 2 presents the results for each cancer’s sensitivity and specificity.

## Discussion

Each of the five cancers were diagnosed with sensitivities of 100% while specificities ranged from 100% for ovarian, CLC, and pancreatic cancers to 96.6% for breast cancer. Tissue of origin was determined with 98% overall efficiency for the MCD. The test is particularly effective at detecting Stage I cancers which is an essential feature of any MCD and is lacking in most other MCD studies. The high tissue of origin makes for minimal downstream testing to confirm the presence of a cancer as one generally does not have to test for more than one cancer type to achieve a diagnosis. The results for breast and lung cancer also support the case for potentially using these specific tests independently as confirmatory tests for cancer following screening mammograms or LD-CT scans respectively.

## Conclusion

An optimal MCED would efficiently detect Stage I disease, have minimal false positives, and would have a high TOO to minimize required downstream testing and biopsies. Nearly 300 samples were tested to diagnose five different cancers (breast, lung, colorectal, ovarian, and pancreatic) relative to apparently healthy normals. The sensitivity for the test was 100% with an overall specificity of 97%. The test detected 100% of Stage I cancers. Cancer patterns did not overlap between cancers such that the cancer identified was the only probable cancer present 98% of the time (TOO of 98%). The SCLC subtype also was differentiated from the NSCLC subtype and presumptive mixed histology samples (both SCLC and NSCLC present) also were identified. For ovarian cancer, the serous subtype, which sometimes contains aggressive cancers, could be differentiated from the mucinous subtype.

This work was supported by Traxxsson, LLC, Saint Louis, MO, USA

## Data Availability

Data in the present study is not available as it contains trade secrets with regard to the biomarkers used.

## Generative AI Statement

The authors declare that no Generative AI was used in the creation of this manuscript.

